# Molecular Epidemiology and Diversity of SARS-CoV-2 in Ethiopia, 2020-2022

**DOI:** 10.1101/2023.01.30.23285174

**Authors:** Abay Sisay, Derek Tshiabuila, Abraham Tesfaye, Gerald Mboowa, Samuel O. Oyola, Sofonias Kifle Tesema, Cheryl Baxter, Darren Martin, Richard Lessells, Houriiyah Tegally, Stephanie van Wyk, Monika Moir, Jennifer Giandhari, Sureshnee Pillay, Lavanya Singh, Yajna Ramphal, Arisha Maharaj, Yusasha Pillay, Akhil Maharaj, Yeshnee Naidoo, Upasana Ramphal, Lucious Chabuka, Eduan Wilkinson, Tulio de Oliveira, Adey Feleke Desta, James E. San

## Abstract

Ethiopia is the second most populous country in Africa and the sixth most affected by COVID-19 on the continent. Despite having experienced five infection waves, >499 000 cases, and ∼7 500 COVID-19-related deaths as of January 2023, there is still no detailed genomic epidemiological report on the introduction and spread of SARS-CoV-2 in Ethiopia. In this study, we reconstructed and elucidated the COVID-19 epidemic dynamics. Specifically, we investigated the introduction, local transmission, ongoing evolution, and spread of SARS-CoV-2 during the first four infection waves using 353 high-quality near-whole genomes sampled in Ethiopia. Our results show that whereas viral introductions seeded the first wave, subsequent waves were seeded by local transmission. The B.1.480 lineage emerged in the first wave and notably remained in circulation even after the emergence of the Alpha variant. The B.1.480 was out-competed by the Delta variant. Notably, Ethiopia’s lack of local sequencing capacity was further limited by sporadic, uneven, and insufficient sampling that limited the incorporation of genomic epidemiology in the epidemic public health response in Ethiopia. These results highlight Ethiopia’s role in SARS-CoV-2 dissemination and the urgent need for balanced, near-real-time genomic sequencing.

## Introduction

Ethiopia is a landlocked country in Eastern Africa [1, 2]. It is further one of the most populous countries on the continent, and following the global trend, has accordingly endured severe economic and healthcare burdens due to the ongoing COVID-19 pandemic [3,4]. On 13 March 2020, the Federal Ministry of Health in Ethiopia reported its first COVID-19 case in Addis Ababa. By February 2022, Ethiopia had recorded approximately 494,760 confirmed cases and 7,572 deaths [5, 6]. Despite various pandemic mitigation strategies implemented by the government (Table S1), the number of COVID-19 cases continued to rise. Ethiopia currently constitutes one of the most severely affected countries on the African continent. On the 8^th^ of April 2020, the Ethiopian government declared a 5-month national state of emergency to control and contain the growth rate of the local epidemic and support mitigation mechanisms [7,8]. These emergency measures included the closing of schools and other academic institutes, restrictions on large gatherings, and implementation of mask mandates. These restrictions were complemented by encouraging social distancing and hand washing using multimedia campaigns [7, 9, 10]. While genetic diversity is a major driver of epidemic dynamics, until now, no detailed molecular epidemiology and genetic diversity analyses of SARS-CoV-2 have been performed in Ethiopia. To address this problem, we analyzed 353 high-quality near complete SARS-CoV-2 genomes sampled in Ethiopia against a globally representative set to characterize the introduction, transmission, and spread of SARS-CoV-2 in Ethiopia.

## Materials and Methods

### Ethical Consideration

This study was reviewed and got approved by Addis Ababa University, College of Health Science Institutional Review Board (IRB) (IRB # 029/20/Lab), IRB of the Department of Medical Laboratory Sciences, Addis Ababa University (reference #- MLS/174/21), IRB office of Addis Ababa City Administration Health Bureau, AAPHREML (Reference #- AAHB/4039/227) and also from Addis Ababa University, College of Natural and Computational Science IRB (IRB #- CNCSDO/604/13/2021), by Yekatit 12 Hospital Medical College (IRB protocol # 75/20) and also reviewed and approved by the Federal Democratic Republic of Ethiopia National Ethics Committee at the Ministry of Science and Higher Education (MoSHE) (IRB #021/246/820/21).

### Study Design, Period, and Setting

We leveraged samples collected from confirmed Ethiopian SARS-CoV-2 cases recorded between July 2020 and February 2022 to produce whole and near-whole genomes. Specifically, whole genome sequencing (WGS) was carried out on a subset of RT-PCR positive samples (N=353) collected during the first four waves of the pandemic in Ethiopia. The genomes can be further classified according to the sampling strategy, i.e., community surveillance (n=170), contacts of confirmed cases (n=78), and suspected cases (symptomatic but yet to be confirmed by laboratory results) (n=105), (Figure S2). Nasopharyngeal (NP) swab specimens were collected using 2ml of Viral Transport Medium (VTM), (China, Miraclean Technology Co., Ltd., www.mantacc.com) from patients in Addis Ababa, Amhara, Oromia, and the Southern Nations, Nationalities, and Peoples Regional States (provinces) of the country. The number of samples selected from each region was based on the epidemic distribution of the diseases across the country (Figure S3). These areas are known to have high population densities and cover almost three-quarters of the country’s inhabitants [11, 12]. Only samples with cycle threshold (Ct) values under 30 were considered viable and submitted for sequencing

### Quality Assurance

We implemented a comprehensive total quality management system to ensure the quality of our sequence data throughout the process (from pre-analytical to post-analytical). The specimens with RT-PCR Ct values <30 were stored in a −80°C freezer at Arsho Advance Medical Laboratories Plc: an ISO 15189 accredited laboratory [13].

### RNA Extraction and Viral Detection

Extraction, purification, and viral detection using RT-PCR were done as per our previous work and respective manufacturer recommendations [14-17]. Viral detection was done at the Ethiopian Public Health Institute (EPHI), AAPHREM center laboratory, Yekatit 12 Hospital, Arsho medical laboratory, and Addis Ababa University.

These were amongst the established RT-PCR laboratories where the majority of COVID-19 testing was performed. SARS-CoV-2 positive samples from the first three waves of the pandemic were shipped to CERI/KRISP (South Africa), and from the fourth wave (103) to ILRI (Kenya) for whole genome sequencing. All biological specimens were transported following a triple packaging biosafety and biosecurity protocol. Samples at CERI/ KRISP were sequenced between 22 September 2021 and 18 October 2022, while samples sent to ILRI were sequenced on March 2022.

### Next-Generation Sequencing of SARS-CoV-2

WGS was performed using either the Illumina or the Oxford Nanopore Technology (ONT) sequencing platforms. Briefly, RNA was extracted on the automated Chemagic 360 system (Perkin Elmer) as per the manufacturer’s instructions. Samples were lysed using proteinase K and bound using silica magnetic beads. Beads were then washed and the RNA was eluted and stored at -80 °C until use. SuperScript IV reverse transcriptase (Life Technologies) was then used, in combination with random hexamer primers, to convert extracted RNA to complementary DNA. Gene-specific multiplex PCR using the ARCTIC protocol was then performed. PCR was used to generate 400 base pair (bp) amplicons, with an overlap of 70 bp to cover the 30 kilobases (kb) SARS-CoV-2 genome. PCR products were purified using AmpureXP purification beads (Beckman Coulter, High Wycombe, UK), and quantification was performed using the Qubit double-strand DNA (dsDNA) High Sensitivity Assay Kit on a Qubit 4.0 instrument (Life Technologies). These libraries were then used for Illumina MiSeq and ONT GridION sequencing.

Illumina MiSeq indexed paired-end libraries were prepared using the Nextera DNA Flex Library Prep Kits (Illumina) as per the manufacturer’s instructions. Tagmented amplicons were barcoded with a unique barcode using the Nextera CD Indexes (Illumina), which allows for library pooling. Prior to pooling, libraries were normalized to 4 nM and pooled, then denatured using sodium acetate. Pooled libraries were then diluted and spiked with 1 % PhiX Control v3 and sequenced using a 500-cycle v2 MiSeq Reagent Kit on the Illumina MiSeq instrument (Illumina, San Diego, CA, USA) [18. ONT Native Barcoding Expansion kits were used for ONT GridION sequencing according to the manufacturer’s protocol. Samples were multiplexed on the FLO-MIN106 flowcells and ran on the GridION X5. The MinKNOW software was used to monitor sequencing performance in real-time [19].

### Metadata Management

Clinical and demographic metadata were collected from the confirmed cases using a structured assessment tool by trained healthcare professionals at the triage of the health facilities and from the community. A medical record number (MRN) was assigned to each patient as a unique anonymous identifier as their names were not linked to the MRNs submitted to sequence repositories such as GISAID, EPI_SET_221214bg doi: <u>10.55876/gis8.221214bg</u>.

### Sequence Alignment, Assembly, and Phylogenetic Analysis

Fastq sequences were assembled using Genome Detective 1.133 (http://www.genomedetective.com) [20] and the Coronavirus tool [21]. The initial assemblies obtained from Genome Detective were visualized and manually edited using Geneious Prime version 2022.2.2 [22,23] to remove low-quality mutations. Nextclade was used to determine the quality control profiles of the sequences. The quality metrics used for the sequences in this study included genome coverage greater than 80%, ensured the removal of unknown frameshifts, stop codon, and clustered mutations. Additionally, submissions with exceedingly high and missing data (N) <3000Ns) were excluded from downstream analyses. Nextclade was further employed to assign clades and lineage classifications to the sequences [24,25]. Sequences that met these quality control thresholds were deposited in the GISAID (https://www.gisaid.org/) database, EPI_SET_221214bg doi: <u>10.55876/gis8.221214bg</u>].

To present a comprehensive analysis of the genomic epidemiology of SARS-CoV-2 in Ethiopia, the genomes generated in this study were combined with a globally representative set of genomes (n= 13589) that were sampled during the same period. All samples from the bordering countries to Ethiopia were included in the analysis to better model the relationship between Ethiopia and its Neighbors and infer cross-border transmission. This did not cause a sampling bias as these countries were already characterized by a low number of sequences. Additionally, all global sequences belonging to the B.1.480 lineage which was predominant in Ethiopia and remained in circulation across the first three waves were included. Together, this resulted in a dataset of 13,942 genomes. Acknowledgment was given to the sequencing laboratories that contributed to the analysis set (EPI_SET_221214bg, DOI: 10.55876/gis8.221214bg).

Sequences were aligned using NextAlign [26] to obtain a good codon alignment of the sequences. A maximum likelihood tree topology was inferred from the resulting alignment in IQTREE 2 [27] using the General Time Reversible model of nucleotide substitution [28] The inferred phylogeny was used along with the associated metadata that was obtained from GISAID, to map discrete geographical locations to each of the tips and infer locations for the internal nodes. This was done using the Mugration package extension of TreeTime [29]. A custom Python script was then used to count the number of discrete changes occurring as we transcended the topology from the root towards the tips. In essence, this provided a crude estimate of the number and timing of viral exchanges (import-export) between Ethiopia and the rest of the world.

### Data Analysis and Visualization

The data were analyzed and visualized using R version 4.2.0 using available packages and custom scripts. The tree was visualized using the R package ggtree version 3.0.4[30].

## Results

### Socio-demographic Characteristics of the Study Participants along with SARS-CoV-2 Variants of Concerns (VOCs)

A total of 1300 nasopharyngeal swabs were collected from four provinces in the country. These samples included 181 (51.3%) specimens obtained from female patients while 172 (48.7%) were sampled from male patients. The patient ages ranged between 2 and 86 years old, although most of the cases were between 21- and 30-year-old patients, with an overall estimated median age of 31.45 (IQR=26-46) years. More than 77 % of the patients presented with mild cases (Table 1). Of these, we discarded 300 due to their high CT values (i.e., > 30). The remaining 1000 samples were subjected to the library preparation and 808 yielded sufficient RNA for WGS following RT-PCR amplification. The WGS resulted in 353 SARS-CoV-2 sequences that passed bioinformatics quality controls. (i.e., with genome coverage greater than 80%, a limited number of private mutations, no clustered mutations, and no misplaced stop codons). Figure S2 illustrates a detailed breakdown of the sample processing. During the study period, a combination of genetically distinct SARS-CoV-2 lineages were identified. The majority of these assemblies (91.7%) were classified as variants of concern (VOC).

**Table 1.**
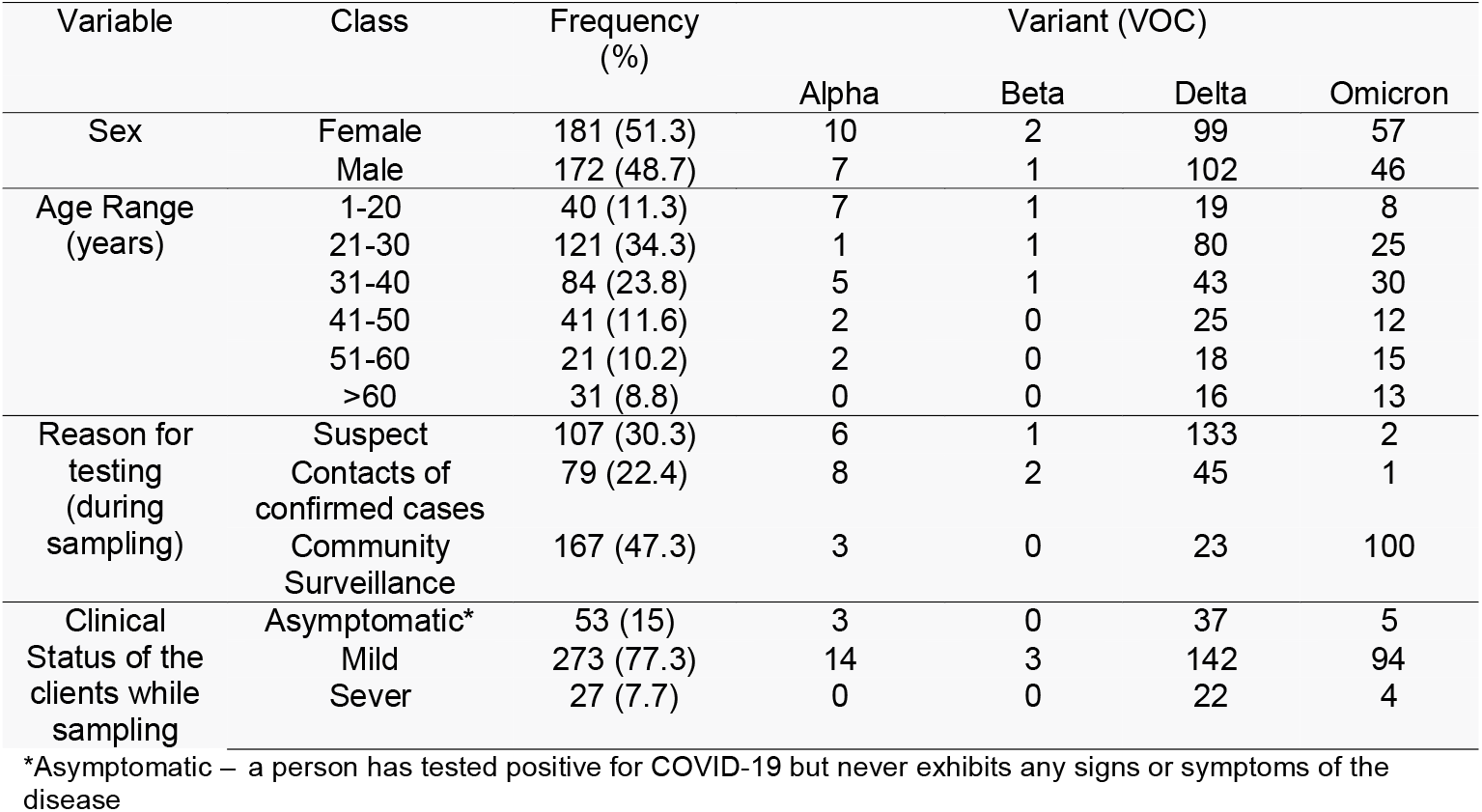
Socio-demographic characteristics of study participants along with VOC of SARS-CoV-2 in Ethiopia, 2020-2022.

| Variable | Class | Frequency (%) | Variant (VOC) |  |  |  |
| --- | --- | --- | --- | --- | --- | --- |
|  |  |  | Alpha | Beta | Delta | Omicron |
| Sex | Female | 181 (51.3) | 10 | 2 | 99 | 57 |
|  | Male | 172 (48.7) | 7 | 1 | 102 | 46 |
| Age Range (years) | 1-20 | 40 (11.3) | 7 | 1 | 19 | 8 |
|  | 21-30 | 121 (34.3) | 1 | 1 | 80 | 25 |
|  | 31-40 | 84 (23.8) | 5 | 1 | 43 | 30 |
|  | 41-50 | 41 (11.6) | 2 | 0 | 25 | 12 |
|  | 51-60 | 21 (10.2) | 2 | 0 | 18 | 15 |
|  | >60 | 31 (8.8) | 0 | 0 | 16 | 13 |
| Reason for testing (during sampling) | Suspect | 107 (30.3) | 6 | 1 | 133 | 2 |
|  | Contacts of confirmed cases | 79 (22.4) | 8 | 2 | 45 | 1 |
|  | Community Surveillance | 167 (47.3) | 3 | 0 | 23 | 100 |
| Clinical Status of the clients while sampling | Asymptomatic* | 53 (15) | 3 | 0 | 37 | 5 |
|  | Mild | 273 (77.3) | 14 | 3 | 142 | 94 |
|  | Sever | 27 (7.7) | 0 | 0 | 22 | 4 |

\*Asymptomatic – a person has tested positive for COVID-19 but never exhibits any signs or symptoms of the disease

### Local Epidemic Dynamics

To illuminate the local epidemic dynamics in Ethiopia, we performed an in-depth study of the 353 high-quality SARS-CoV-2 genomes (Figure 1). The study period corresponds with the first four COVID-19 waves experienced in the country. The first wave occurred between August and September 2020, the second between February and March 2021, and the third occurred between July and October 2021. Each wave was characterized by higher rates of transmission and fatality than previous waves (Figure 1A). Notably, the third wave was more severe and lasted longer than the two previous waves. The fourth wave began with an uptick in cases in December 2021 and continued to February 2022. The wave was characterized by the highest rates of community transmission yet observed (Figure 1A). However, compared to the global trend at the time, it resulted in fewer deaths.

**Figure 1:**
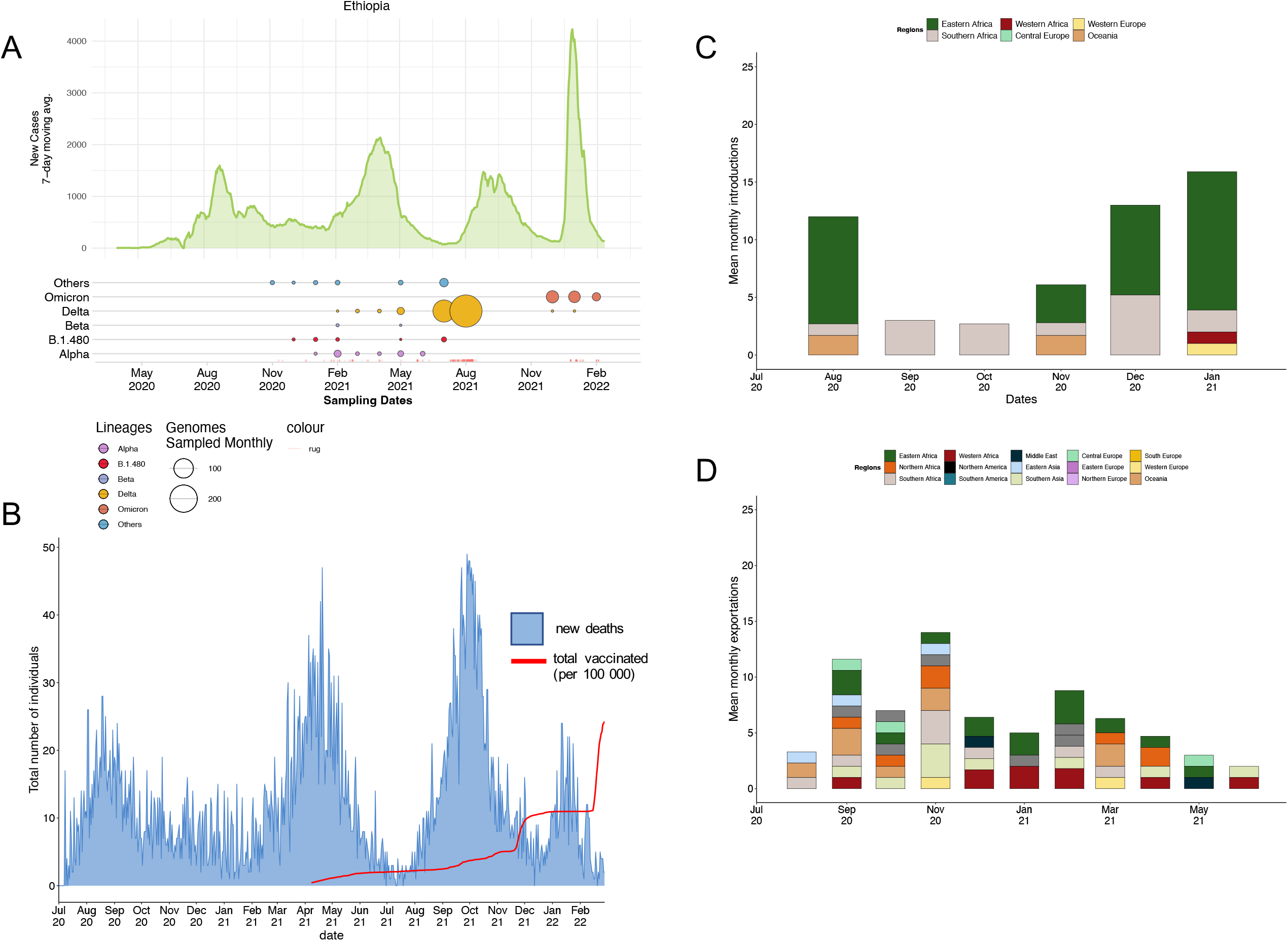
SARS-CoV-2 epidemiology and variant turnover in Ethiopia: (a) 7-day rolling average for the number of new SARS-CoV-2 cases. Others, represented in blue, comprise mostly B lineages including B, B.1, and B.1.1 lineages. The size of the dot is indicative of the variant proportion in the population. (b) The proportion of individuals that have received at least one vaccine dose in Ethiopia compared to the number of new deaths that occurred. (c) Summary of the mean number of detected introductions into Ethiopia binned by month for different regions across the dataset. (d) Summary of the mean number of detected exportations out of Ethiopia to different regions binned by month.

Across the four waves, the peak number of new cases remained relatively consistent with the first and third waves reflecting a 7-day rolling average of approximately 1500 individuals. The second wave had a slightly higher 7-day rolling average of 2000 individuals whilst the fourth had the highest at over 4000 individuals (Figure 1A). The number of new deaths across each wave had a similar trend to the average number of new cases with the second and third waves having the highest number of new deaths at approximately 50 individuals per day (Figure 1B). Vaccine rollout commenced in April 2021 in Ethiopia and by February 2022, approximately 25% of the population had received at least one dose of Oxford-AstraZeneca or Sinopharm COVID-19 vaccines (Figure 1B).

### Phylogenetic reconstruction and Variant Detection in Ethiopia

Ancestral state reconstruction showed that viral imports into Ethiopia occurred between August 2020 and January 2021, with the majority of importation events occurring in August 2020 [n=123], December 2020 [n=130], and January 2021 [n=169] (Figure 1C). In comparison, September and November 2020 corresponded with the highest numbers of inferred SARS-CoV-2 exports from Ethiopia (n=109 and n =137, respectively; Figure 1D, Figure S4). The majority of inferred importation events were from Eastern Africa [n=324] origins, whereas most of the inferred exportation events were generally attributed to Western European and Asian origins (n=20 and n=141 respectively). Overall, we identified 570 importation and 446 exportation events during the study period.

Our early sequences sampled between November and December 2020 represented two lineages, B.1/B.1.1 and B.1.480, with most of the sequences classified as B.1/B.1.1. These two lineages continued to circulate throughout the first quarter of 2021 (January to March). The B.1.480 lineage was thought to have originated in Ethiopia. In our study, 21 genome assemblies were assigned to this emerging lineage. (EPI_SET_230126dy, doi: 10.55876/gis8.230126dy). Phylogenetic inference supported that this lineage was a monophyletic clade. Despite its persistence in Ethiopian populations for an extended period, this variant did not disseminate globally as was observed for other VOC and ultimately only constituted only 473 GISAID submissions during the progression of the pandemic, EPI_SET_230126vf, doi: 10.55876/gis8.230126vf. Nevertheless, according to GISAID submissions, this lineage was detected in neighboring countries such as Kenya, Somalia, Djibouti, and Guinea-Bissau. This lineage was further disseminated internationally where it was detected in Australia, the United States of America, Canada, Japan, Israel, Hong Kong, and South Korea. This period was followed by the detection and rise to dominance of VOCs in the country.

In January 2021, the first case of the Alpha variant was detected. Within a span of one month, it had reached equal prevalence to B.1/B.1.1. The B.1.1 continued to circulate in low prevalence with the cases detected until May 2021. Alpha was however short-lived as the Delta variant emerged. The Delta variant was first detected in February 2021 and continued circulating in low prevalence until May 2021. Thereafter it outcompeted all other lineages and variants in circulation to become the most prevalent variant (Figure S5). The predominant Delta variant sub lineages where AY.120 (116 sequences, 46.4%) and B.1.617.2 (50 sequences, 20%). The Delta variant seeded the 3^rd^ wave in Ethiopia. Although no sequence data was available from September to December 2021, the majority of the cases sampled and sequenced from January 2022 were attributed to the Omicron lineage (Fig 1A & 2A). Over the course of the study period, we identified 37 distinct lineages in circulation (Fig 2 A, Fig 2B, Figure S6).

**Figure 2:**
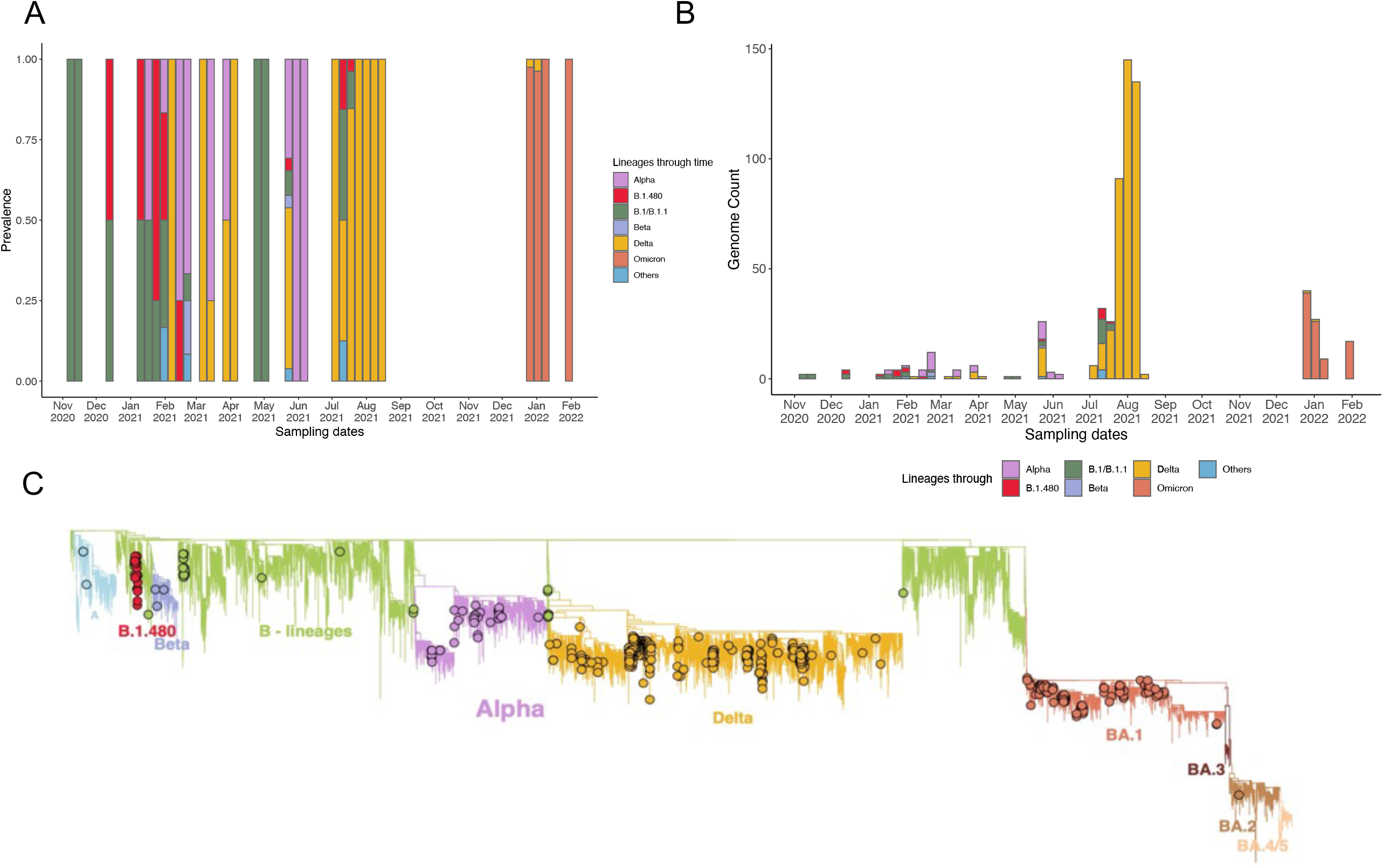
SARS-CoV-2 lineage diversity in Ethiopia. (A) Proportions of SARS-CoV-2 lineages, classified by Pangolin nomenclature, circulating between June 2020 and February 2022 in Ethiopia. (B) Absolute counts of SARS-CoV-2 genomes throughout the study period. Only VOCs and major lineages are listed in the legend. (C) Maximum-Likelihood phylogeny of 353 SARS-CoV-2 genomes. Branches are colored by lineage.

The phylogenetic reconstruction revealed close relatedness (not substantially observed levels of divergence) among sequences assigned to the same lineages. However, this observation did not hold for samples assigned to the Delta variant (Figure 2C). Sequences attributed to the Delta variant showed greater variation in sequence composition. Moreover, higher levels of phylogenetic clusters, associated with community transmission, were observed during the study period. Phylogenetic clusters and associated community transmission events were also observed for the Omicron lineage (Figure 2C).

## Discussion

In this study, we investigated the introduction of SARS-CoV-2 viral lineages, local transmission, and its evolution within Ethiopia. This study represents the first of its kind as no other molecular epidemiology investigation has been conducted for this country and provided an overview of the pandemic progression relative to a global context. Our results show that SARS-CoV-2 was introduced into Ethiopia on multiple occasions, with most introductions originating from Eastern African countries. These introductions included 37 distinct SARS-CoV-2 lineages and four VOCs, namely Alpha, Beta, Delta, and Omicron lineages. Of the introduced lineages the B1.480 persisted and remained in circulation from December 2020 to August of 2021.

Our findings show that as of February 2022, Ethiopia had experienced four distinct COVID-19 waves. Although various preventive strategies such as the mandatory wearing of masks, and media awareness campaigns were adopted together with a mandatory 14-day self-quarantine regulation for all international travelers arriving in the country [5], Ethiopia remained vulnerable to SARS-CoV-2 introductions. Our results showed that despite preventative measures viral introduction events continued to occur and were followed by community transmission. This in turn facilitated a continued increase in case numbers. This could be attributed to the partial restriction of public transport which is an import mode of SARS-CoV-2 transmission [5, 31]. During lockdowns, the operation of economic activities did not cease as they were deemed necessary to support the already fragile economic and socioeconomic situation of the country. This made lockdowns ineffective in preventing local transmission and epidemic growth. By 30 April 2021, the total number of cases and deaths had surpassed 257442 and 3688, respectively [1, 3], and has currently increased to >499000 and 7500 deaths.

Our findings show that Ethiopia experienced poor sampling mostly limited to Addis Ababa. Temporal sampling was also hampered by periods of limited sampling and, lack of local sequencing capacity resulting in substantial delays that in turn may cause the degradation of samples during transit. Yet, an in-depth understanding of the COVID-19 pandemic progression in Ethiopia remains of paramount importance to aid the navigation of ongoing and future pandemics within these regions [32]. Indeed, Ethiopia has had several viral outbreaks in recent history, in addition to the ongoing COVID-19 pandemic. Some of the notable outbreaks include Measles, and mosquito-borne illnesses such as Yellow fever, Chikungunya, and Malaria [32, 33]. Thus, scaling up of local and regional pathogen genomic sequencing capacity and a robust pathogen genomic surveillance infrastructure is thus required to shorten the turnaround time and to allow for early public health response [34]. Additionally, during the onset of the pandemic, a survey on outbreak readiness highlighted a dire need for not only pathogen surveillance in Ethiopia but Ethiopia further scored only 52% on the Ready Score index, which shows that the nation still has progress to be made to increase case tracing and identification, increase healthcare and testing facilities, provide clear operational guidelines on preventive measures across various organizations, businesses, and community settings, as well as proactive steps to maintain life during social and economic lockdowns[3, 35]. Ethiopia, as with many other African countries, has limited resources to deal with these outbreaks, and often depends on international aid to help control and prevent these diseases [10, 36, 37].

Over the course of four waves, we estimated 570 introductions and 446 exportation events occurred. This is consistent with the relatively small number of lineages identified during the study period, although could also be attributed to under and sparse sampling that likely omitted lineages circulating at low prevalence. The short generation time associated with RNA viruses could also imply that viruses circulating during the period preceding the emergence of Omicron that became extinct would not have been detected. The phylogenetic inference and that of the geographical distribution of the Ethiopian-emerging B.1.480 lineage suggest that Ethiopia serves as a source for viral introductions not only to neighboring countries but also further abroad. Ethiopia serves as a travel hub for this African region connecting this region in Africa to neighboring countries and further abroad. Possible disease dissemination was most likely a result of travel to and from Ethiopia via air, land, and waterways [37, 38]. Ethiopia is strategically located on the Red Sea, making it an important transit point for goods moving between Africa, the Middle East, and Asia. The main international airport in Ethiopia is Addis Ababa Bole International Airport facilitates high travel volumes which most likely contributed to viral amplification in the country followed by Ethiopian-originating viral exportation events. Therefore, concerted, collaborative action is imperative to achieve pandemic mitigation.

We observed a reduction in the number of new deaths reported during the fourth wave. This is consistent with the immunity acquired protection from a large number of infections from the preceding waves and increases in the total number of vaccinated individuals. A sero-prevalence study among healthcare workers revealed a seropositivity rate of 39.6% [39] while another study estimated the prevalence rate at over 50% [40]. However, only about 0.4% of the population has reportedly been infected with COVID-19: much fewer than the estimated sero-prevalence rates estimated for the population. Therefore, taken together these statistics suggest that case numbers may remain widely underreported. Reduced testing further hinders effective genome surveillance as fewer samples were available to be subjected to whole genome analyses and influences the accuracy of epidemiological conclusions drawn.

In comparison to other countries, Ethiopia has very low vaccination coverage. As of December 2021, at least 1 000 000 Ethiopians had been vaccinated. It is important to note that COVID-19 vaccines in Ethiopia were prioritized based on criteria such as occupation, age, medical condition, people that work in aggregate settings, the elderly, and people with comorbidities known to increase the risk of adverse COVID-19 outcomes [41]. Moreover, priority was given to those deemed as frontline medical staff, followed by non-frontline medical staff. This strategy was effective in slowing the evolutionary rate of the virus as SARS-CoV-2 is prone to evolution in immunosuppressed individuals experiencing chronic infection [42-44]. Vaccination has also been shown to decrease the overall SARS-CoV-2 infection rate from 9.0% (95% CrI: 8.4% – 9.4%) without vaccination to 4.6% (95% CrI: 4.3% – 5.0%) with the highest relative reduction noted among individuals aged 65 and above. Adverse outcomes, such as deaths, are reduced by approximately 69.3% (95% CrI: 65.5%-73.1%) with lowered vaccination coverage [45].

Analysis of genomes and lineages across the study period highlighted the changes in the population dynamics of the viral lineages circulating in Ethiopia. The B.1 and B.1.1 lineages dominated the first wave, persisted throughout the next two waves, and were characterized by the well-known D614G mutation [46]. This mutation has been linked to increased viral replication and transmissibility [38, 46]. Subsequent waves were dominated by the VOCs Alpha, Beta, Delta, and Omicron. These VOCs evolved to evade overcome host immunity from infection with ancestral viruses and vaccination. By doing so these newly emerging variants fueled renewed infectious waves and were often associated with increased transmissibility compared to their genetic predecessors [47-49]. The Omicron variant (specifically, lineage BA.1.1) had a reproduction number four times higher than Delta. It is considered the fifth variant of concern first detected in November/December 2021 [50-53].

One important lineage in the context of Ethiopia was the B.1.480, a distinct B lineage, which emerged during the second wave. This variant was responsible for 2.77% of the sampled cases. Notably, this variant was previously detected in passengers originating from not only Ethiopia but also with those originating from European countries such as the United Kingdom and France, as well as from travelers from United Arab Emeritus [50-53]). Unlike other VOCs, this lineage did not reach global transmission and has only 473 submissions to the GISAID platform, however, it did persist in Ethiopian populations for extended periods of time. Other worrisome mutations carried by this lineage included the Spike mutation M1229I and D614G [46,47,49,54].

We observed early introductions of the Delta variant as early as February 2021. To rule out contamination as possibly a source of these sequences, we examine mutational patterns within these sequences, nucleotide distributions in the sorted, indexed and mapped bam files, and the timing of sequencing. Considering the timing of sequencing and that these samples where sequenced along with several samples that were also attributed to the Delta variant, contamination as a possible source could not be entirely ruled out. However, we note that these sequences conform to molecular clock estimations of the emergence of Delta globally estimated around 19 October 2020 with an earliest possible date estimated as 06 September 2020 [55], Figure S5. A similar pattern of early emergence and slow rise to dominance is observed in several countries across the continent [34] and, in a more detailed study in Neighboring Kenya [56].

In conclusion, the changing nature of the pandemic over time in Ethiopia has been driven by lineage turnovers that, while dominated by VOCs and B.1.480, include other lineages (A, B.1, B.1.1). Results from this study, and that of others (ref listed above), showed that despite pandemic mitigation strategies implemented by the country, Ethiopia remained vulnerable to viral introduction events and served as a source of viral introductions internationally. The COVID-19 pandemic is still ongoing and the evolution of the virus is continuing to yield new variants some of which will likely reflect altered viral behavior such as increased transmissibility, immune evasiveness, or altered pathogenicity. It is therefore crucial that ongoing genomic surveillance be strengthened and conducted to present real-time data generation to determine the geographic distribution of these variants and enable the implementation of appropriate disease mitigation strategies.

## Data Availability

All the generated sequences used for this manuscript have been deposited in the GISAID sequence database and are purely publicly available. However its subject to the terms and conditions of the GISAID database https://www.gisaid.org/GISAID Identifier: EPI_SET_221214bg DOI: 10.55876/gis8.221214bg.

https://gisaid.org/epi_set_221214bg

https://doi.org/10.55876/gis8.221214bg

## Acronyms /abbreviations

AAPHREML: Addis Ababa public health research and emergency management laboratory
AAU: Addis Ababa University
ACDC: Africa center for disease prevention and control
CERI: Centre for Epidemic Response and Innovation
CI: Confidence Interval
CT: Cycle Threshold
GISAID: Global Initiative on Sharing All Influenza Data
KRISP: KwaZulu-Natal Research Innovation and Sequencing Platform
NAAT: Nucleic Acid Amplification Tests
PCR: Polymerase Chain Reaction
RNA: Ribose Nucleic Acid
RT-PCR: Reverse Transcriptase Polymerase Chain Reaction
SARS-CoV-2: severe acute respiratory syndrome novel corona virus-2
VOCs: Variants of Concern
VTM: Virus Transport Medium
WHO: World Health Organization
SDG: sustainable development goal
COVID-19: Coronavirus disease 2019

## Author Contributions

AS and AFD conceived and designed the experiments; AS, DT and SEJ performed the data analysis and experiments; AS, DT, AFD and SEJ analyzed the data; GM, SO, SKT, CB, DM, RL, HT, MM, SvW, EW, TDO contributed materials/analysis tools; AT, AFD and SEM supervise the work; AS, DT, SvW and SEJ wrote the paper. All authors read the final version of the manuscript.

## Informed Consent Statement

All participants gave written informed consent for participation and publication.

## Data availability

All the generated sequences used for this manuscript have been deposited in the GISAID sequence database and are purely publicly available. However, it’s subject to the terms and conditions of the GISAID database, https://www.gisaid.org/, GISAID Identifier: EPI_SET_221214bg, DOI: <u>10.55876/gis8.221214bg</u>.

## Acknowledgment

We would like to acknowledge Addis Ababa University for giving us the opportunity and financing the research. This work is realized with the generous financial and technical support of Africa CDC, PGI

We are very grateful and acknowledge the CERI and KRISP (South Africa) & ILRI (Kenya) for supporting this work by performing the whole genomic sequencing.

We also wish to extend our gratitude to Arsho Medical Laboratories for supporting this research by giving us a -80 ^Oc^ refrigerator for centrally storing our specimens.

Last but not least we are very glad and thankful to Retina Pharmaceutical for supporting this work by availing required pharmaceutical inquiries.

---

**Table S1.** Summary of the COVID-19 timeline and major events following the first confirmed cases in Ethiopia

| Timeline | Major events | Remark |
| --- | --- | --- |
| December 2019/January 2020 | Wuhan City, Hubei Province, China, which was reported to the WHO on 31 December 2019. Wuhan, China, was reported in January 2020 |  |
| 27 January 2020 | Prior to the WHO's declaration of a global pandemic, the federal democratic republic of Ethiopia Council of Ministers 'activated' a National Public Health Emergency Preparedness Centre, EOC and began preparations to deal with a potential outbreak of Covid-19 |  |
| 30 January 2020 | the Director-General of the WHO declared COVID-19 a Public Health Emergency of International Concern |  |
| 7 February 2020 | Ethiopia set up the national Influenza and Arboviruses reference laboratory as the first COVID-19 testing laboratory in the country with the support from WHO and ACDC. |  |
| March 11, 2020- | WHO declared COVID-19 as a Pandemic |  |
| March 13, 2020,<br>*** | On 12 March 2020, a week after entering the country from Burkina Faso, a 48-year-old Japanese national presented **himself** at a public health center in the capital city, Addis Ababa, and was diagnosed as having COVID-19. Since Ethiopia did not have the capacity to test for COVID-19 at the onset. Thus, samples were initially transported for testing to the National Institute for Communicable Diseases (NICD) Laboratory in South Africa |  |
| 15 March 2020 | The second confirmed case from Ethiopian nationalities (three additional )cases of the COVID-19 were reported |  |
| 20 March 2020 | Anyone entering the country/international travelers/ should stay in quarantine for up to 14 days at their costs |  |
| 23 March 2020 | Ethiopia closed all land borders and deployed security forces to halt the movement of people along the borders (National lockdown....) |  |
| 24 March 2020 | • banned all public gatherings and sports events<br>• The closure of bars and clubs<br>• all federal employees were to work from home, except those designated by each ministry and federal agency as essential workers |  |
| 05 April 2020 | The First COVID-19-related confirmed death |  |
| 8 April 2020 | A state of emergency was declared giving authorities across-the-board powers to battle the disease |  |
| 06 March 2021<br>*\*after a year of pathogen occurrence* | The first genome sequence of SARS-CoV-2 sampled from Ethiopia was submitted to the GISAID database |  |
| 13March, 2021 | First vaccination started |  |

\*\*\* Form 13 March-Mid April 2020-Sample were tested by referral to NICD, the country had no RT-PCR testing center during this time

**Figure S2.**
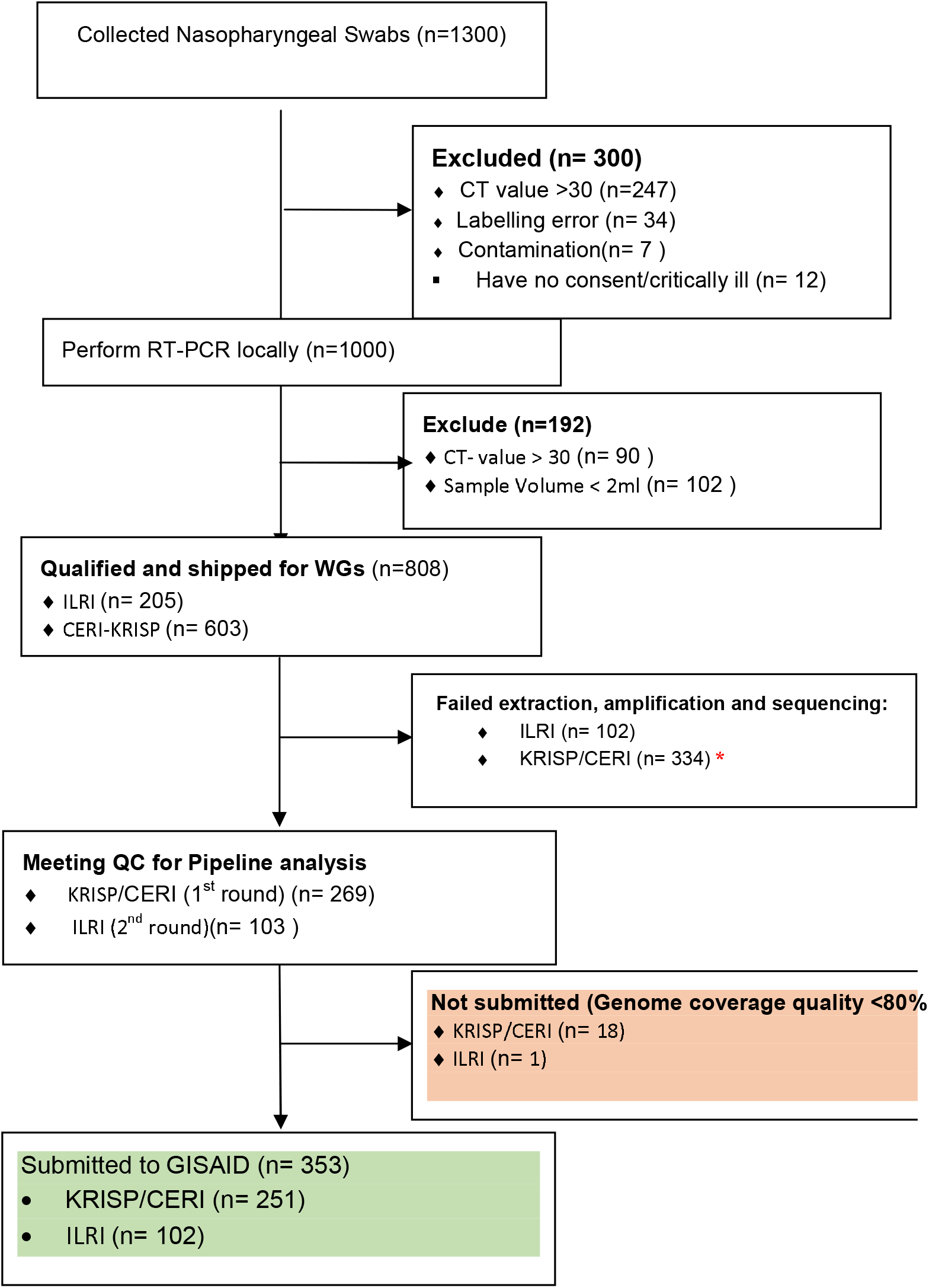
Consortium Diagram of Sampling for Molecular Epidemiology in Ethiopia *because of the old specimen, long shipping time, backlog seq., temp not maintained …

**Figure S3a.**
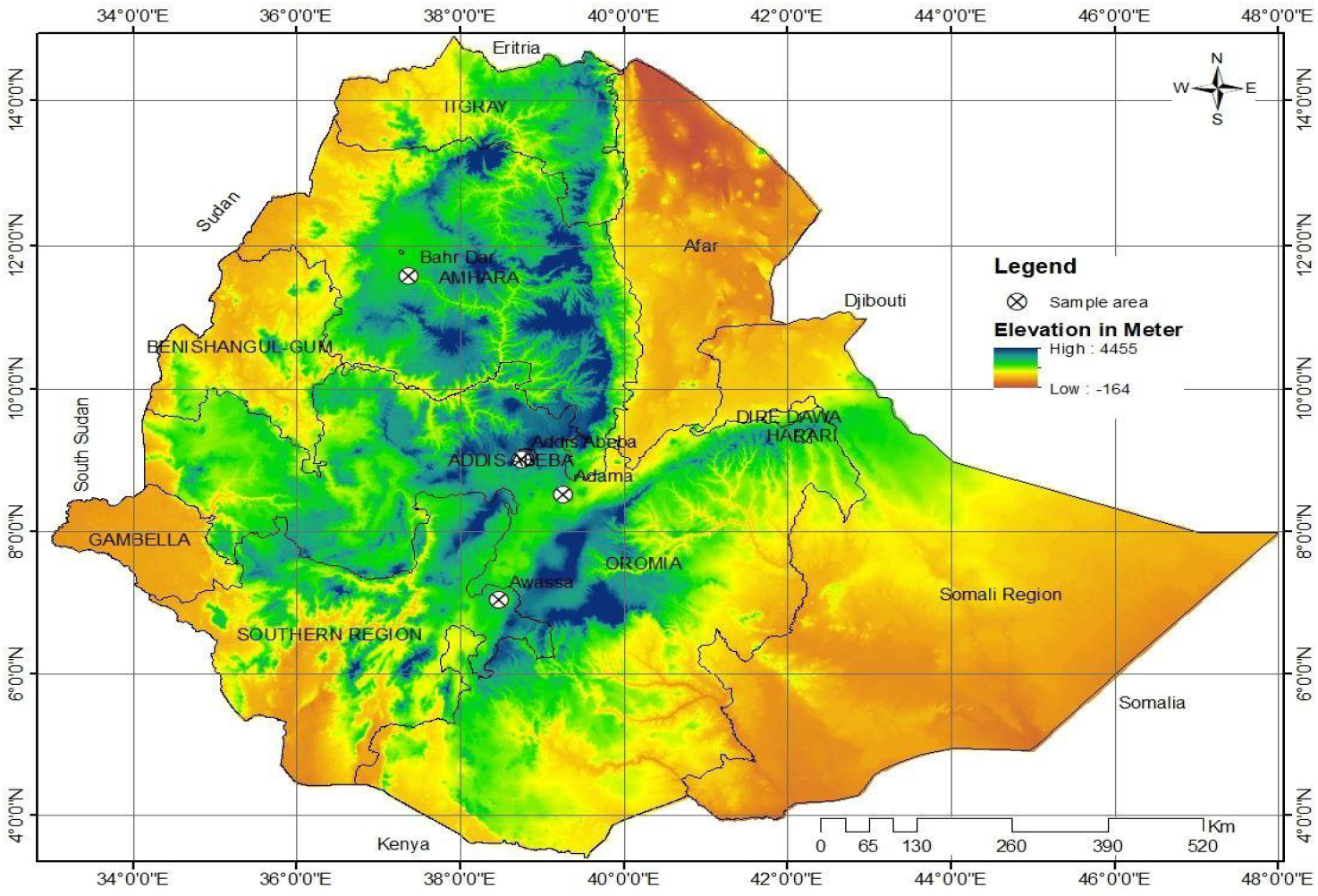
Sampling area for Molecular Epidemiology and Diversity of SARS-CoV-2 in Ethiopia

**Table S3b:** SSequence proportion of the regional government

| Name of Regional | Proportion of sequencing (%) |
| --- | --- |
| Addis Ababa | 80% |
| Oromia | 8% |
| Amhara | 5% |
| SNNPR and Sidama | 5% |
| Others | 2% |

**Figure S 4.**
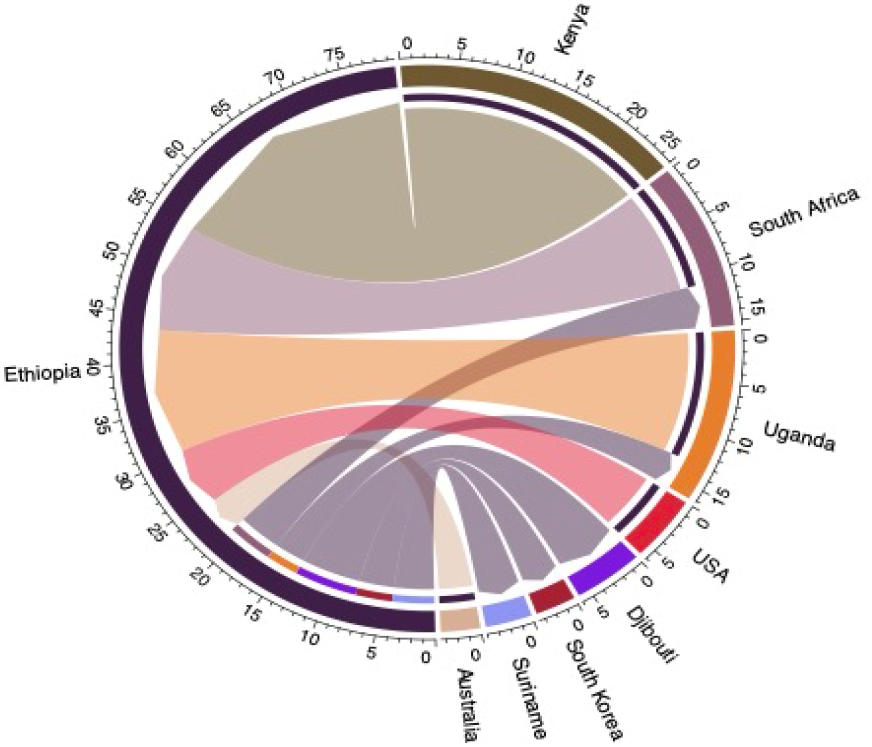
Major SARS-CoV-2 lineages of detected imported and exported Ethiopia between June 2020 and February 2022

**Figure S 5.**
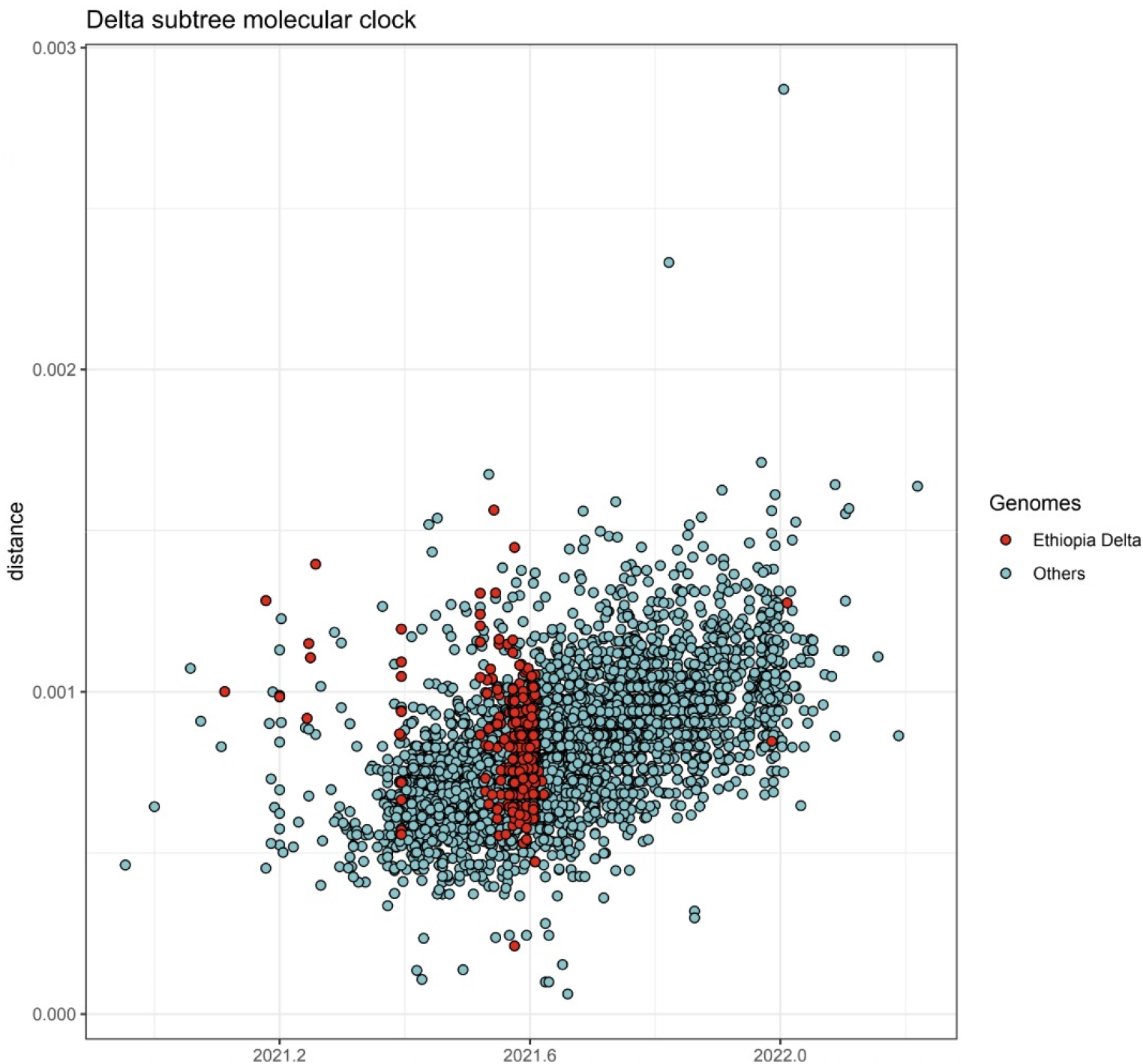
SARS-CoV-2 Delta VOC sub tree molecular clock, Ethiopia.

**Figure S 5.**
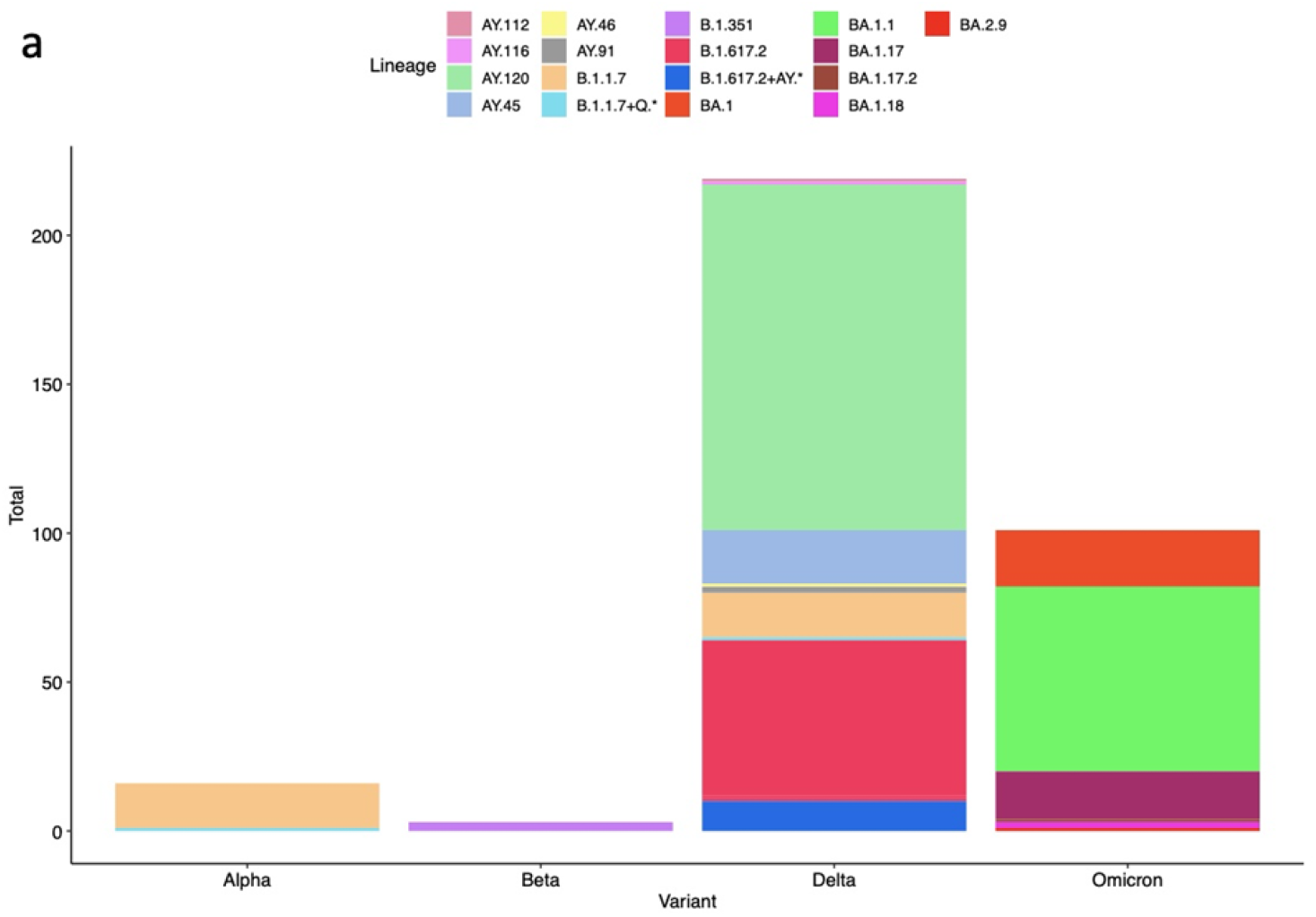
Major SARS-CoV-2 lineages circulating in Ethiopia between June 2020 and February 2022

